# Prognostic significance of BMI after PCI treatment in ST-elevation myocardial infarction A cohort study from the Swedish Coronary Angiography and Angioplasty Registry

**DOI:** 10.1101/2021.01.12.21249477

**Authors:** Shabbar Jamaly, Björn Redfors, Elmir Omerovic, Lena Carlsson, Kristjan Karason

**Author notes:** **Address for correspondence:** Dr Shabbar Jamaly, Department of Cardiology, Sahlgrenska University Hospital 41345 Gothenburg Sweden. **CLINICAL TRIAL REGISTRATION:** NCT02311231.

## Abstract

**Background:** Obesity along with clustering of cardiovascular risk factors is a promoter for coronary artery disease. On the other hand, a high BMI appears to exert a protective effect with respect to outcomes after a coronary artery event, termed the obesity paradox.

**Methods:** The Swedish Coronary and Angiography and Angioplasty registry (SCAAR) collects information on all patients who undergo percutaneous coronary intervention (PCI) for ST-elevation myocardial infarction (STEMI) in Sweden along with demographic- and procedure-related data. We studied the predictability of four categories of BMI for 1-year all-cause mortality in people with STEMI undergoing PCI.

**Results:** Among 25,384 patients, mean (SD) age 67.7 (12.1) years and 71.1% male, who underwent PCI for STEMI a total of 5,529 (21.8%) died within one year. Using normal-weight (BMI 18.5-24.9 kg/m^2^) as a reference, subjects with obesity (BMI ≥30 kg/m^2^) had a low 1-year all-cause mortality risk in unadjusted analysis, HR 0.59 (95% CI 0.53– 0.67). However, after adjustment for age, sex and other covariates the difference became non-significant, HR 0.88 (95% CI: 0.75-1.02). Patients with overweight (BMI 25.0-29.9 kg/m^2^) had the lowest 1-year mortality risk in analysis adjusted for age, sex and other covariates, HR 0.87 (95% CI 0.79-0.95), whereas those with underweight (BMI <18.5 kg/m^2^) had the highest mortality in both unadjusted HR 2.22 (95% CI 1.69–2.92) and adjusted analysis, HR 1.72 (95% CI: 1.31-2.26).

**Conclusion:** The protective effect of obesity with respect to 1-year mortality after coronary intervention became non-significant after adjusting for age, sex and relevant co-variates. Instead, overweight people displayed the lowest risk and underweight individuals the highest risk for adjusted all-cause mortality

## Introduction

Obesity, together with associated clustering of cardiovascular risk factors, such as hypertension (1) dyslipidemia (2) and diabetes (3), is a strong promoter for cardiovascular disease morbidity and mortality (4-6). Weight control is considered to be of fundamental importance in primary prevention aimed at reducing the overall incidence of cardiovascular disease (7) and is also targeted in secondary preventive programs intended to improve outcome in patients with established cardiovascular disease (8, 9).

Still, a certain hesitancy has arisen concerning the beneficial effects of weight loss as a secondary prevention practice, since several epidemiologic have suggested that obesity may be protective in patients with coronary artery disease undergoing intervention (10-12). The apparent favorable effect of obesity on outcomes after coronary interventions, known as the “obesity paradox”, has generated a substantial amount of controversy (13). A protective effect of excess body fat is somewhat counter-intuitive and a mechanism involving reverse causality has been suggested (14). Also, body fat distribution, cardiorespiratory fitness and unintentional weight loss could constitute confounders accounting for the somewhat paradoxical relationship between obesity and prognosis after coronary intervention (15).

The aim of the present study was to evaluate the relationship between body fatness, divided up as four different body mass index (BMI) categories, and mortality in a large Swedish population undergoing percutaneous coronary intervention (PCI) due to an ST-elevation infarction (STEMI).

## Patients and methods

### Patient population

The Swedish Coronary and Angiography and Angioplasty Registry (SCAAR) was established in 1992 and contains information about all coronary angiographies and percutaneous coronary interventions (PCI) (formerly known as angioplasty with stent) (10). Each catheterization procedure is described with approximately 50 angiography and 200 PCI variables, including both demographic and procedure-related data. The registry is financed by the Swedish government and the Association of Local Authorities and Regions and is supported by the Swedish Heart Association, the National board of Health and Welfare and the Swedish Heart and Lung Foundatioion

All consecutive patients undergoing PCI for ST-elevation myocardial infarction (STEMI) in Sweden between January 1^st^ 2011 and May 31^st^ 2018 were included in the study. STEMI was defined according to the European Society of Cardiology criteria (16) as a condition when there is evidence of myocardial injury defined as a dynamic change in cardiac troponin values with at least one value above the 99^th^ percentile upper reference limit, or a persistent chest discomfort suggestive of myocardial ischemia, along with an ST-segment elevation in at least two contiguous leads. Body weight and height, either measured or self-reported, were entered in the register at the time of the intervention. BMI, as a measure of nutritional status, was calculated as the weight in kilograms divided by the square of the height in meters. Other patient characteristics and information on co-morbidities were imported into the register from medical records. All patients admitted to the cardiac care unit are informed both, verbally and in writing about their participation in the registry. The regional ethical review board in Gothenburg approved the study. The investigation conforms with the principles outlined in the Declaration of Helsinki.

### Outcome measures

The primary endpoints were mortality rates at 30 days and at 1 year. All Swedish citizens have a specific personal identity number that is recorded in connection with all health care contacts and makes it feasible to follow how the Swedish population interacts with the health care system. The SCAAR registry obtains data on patients’ vital status from the Swedish Cause of Death Register, which originates from 1952 and includes the cause of mortality for all of citizens registered in Sweden at the time of their death (17).

### Statistics

Statistical analyses were performed with SAS 9.4 statistical software packages (SAS Institute, Cary, NC). Study participants were divided into four categories according to their nutritional status as recommended by the World Health Origination (18): Underweight (BMI <18.5 kg/m^2^), normal weight (BMI 18.5-24.9 kg/m^2^), overweight (BMI 25.0-29.9 kg/m^2^) and obese (BMI ≥30.0 kg/m^2^). Data are presented for the total study population and for each BMI group separately as means and standard deviations, medians and interquartile ranges, or numbers and percentages. Comparisons between groups at baseline based on a complete case analysis were performed with analysis of variance (ANOVA) for normally distributed numeric data, Chi-Square test for categorical data and Kruskal-Wallis rank sum test for non-normally distributed data. The number of complete cases with data for each variable is given as the dominators in **Table 1**.

**Table 1.** Baseline characteristics for the total study group and for the different BMI categories separately*

| Characteristics | Total Group<br>(n=25384) | Underweight<br>(n=244) | Normal weight<br>(n=8488) | Overweight<br>(n=11132) | Obese<br>(n=5520) | P-value |
| --- | --- | --- | --- | --- | --- | --- |
| Age (years) | 67.7 ± 12.1 | 74.8 ± 11.6 | 71.1 ± 12.0 | 67.1 ± 11.5 | 63.6 ± 11.7 | <0.001 |
| Weight (kg) | 81.2 ± 16.0 | 48.2 ± 5.9 | 68.4 ± 9.2 | 82.7 ± 9.5 | 99.7 ± 14.5 | <0.001 |
| Height (cm) | 172.9 ± 9.7 | 167.0 ± 8.9 | 172.2 ± 9.3 | 173.9 ± 8.9 | 172.3 ± 10.6 | <0.001 |
| Body Mass Index (kg/m<sup>2</sup>) | 27.2 ± 6.1 | 17.3 ± 1.1 | 23.0 ± 1.6 | 27.3 ± 1.4 | 33.8 ± 9.4 | <0.001 |
| Female sex | 28.9% (7345/25384) | 68.9% (168/244) | 33.8% (2870/8488) | 23.4% (2606/11132) | 30.8% (1701/5520) | <0.001 |
| Hypertension | 49.0% (12221/24952) | 43.8% (103/235) | 43.9% (3648/8309) | 48.6% (5330/10973) | 57.8% (3140/5435) | <0.001 |
| Hyperlipidemia | 25.5% (6325/24792) | 19.0% (44/231) | 21.5% (1780/8266) | 25.8% (2810/10899) | 31.3% (1691/5396) | <0.001 |
| Diabetes mellitus | 16.2% (4095/25218) | 7.5% (18/241) | 11.3% (955/8427) | 15.0% (1666/11072) | 26.6% (1456/5478) | <0.001 |
| Insulin-treated diabetes mellitus | 7.0% (1756/25169) | 3.7% (9/241) | 4.7% (396/8413) | 6.0% (666/11057) | 12.6% (685/5458) | <0.001 |
| S-creatinine (μmol/L) | 92.4 ± 55.1 | 89.2 ± 65.8 | 92.4 ± 55.7 | 91.3 ± 50.9 | 92.0 ± 53.0 | <0.001 |
| eGFR <30ml/min/1.75m<sup>2</sup> | 1445 ± 5.7 | 24 ± 9.8 | 558 ± 6.6 | 576 ± 5.2 | 287 ± 5.2 | <0.001 |
| Previous renal failure | 2.5% (640/25384) | 2.9% (7/244) | 2.7% (227/8488) | 2.1% (237/11132) | 3.1% (169/5520) | <0.003 |
| Previous dialysis | 0.4% (107/25384) | 0.4% (1/244) | 0.4% (37/8488) | 0.4% (43/11132) | 0.5% (26/5520) | 0.88 |
| Current smoker | 28.6% (6880/24033) | 41.4% (94/227) | 29.9% (2388/7990) | 27.0% (2857/10577) | 29.4% (1541/5239) | <0.001 |
| Previous COPD | 5.3% (1355/25384) | 21.3% (52/244) | 6.4% (546/8488) | 4.1% (456/11132) | 5.5% (301/5520) | <0.001 |
| Previous cancer diagnosis | 2.3% (590/25384) | 3.3% (8/244) | 2.6% (222/8488) | 2.2% (250/11132) | 2.0% (110/5520) | 0.07 |
| Previous dementia | 0.4% (96/25384) | 1.6% (4/244) | 0.4% (38/8488) | 0.4% (43/11132) | 0.2% (11/5520) | <0.001 |
| Previous heart failure | 4.4% (1120/25384) | 6.6% (16/244) | 4.7% (395/8488) | 3.7% (414/11132) | 5.3% (295/5520) | <0.001 |
| Previous myocardial infarction | 13.0% (3296/25384) | 11.9% (29/244) | 12.6% (1073/8488) | 12.6% (1402/11132) | 14.3% (792/5520) | <0.02 |
| Previous PCI | 10.8% (2750/25384) | 7.4% (18/244) | 9.7% (827/8488) | 11.0% (1220/11132) | 12.4% (685/5520) | <0.001 |
| Previous CABG | 2.9% (747/25384) | 1.2% (3/244) | 2.5% (214/8488) | 3.0% (334/11132) | 3.6% (196/5520) | 0.002 |
| Previous stroke | 31.9% (7661/24033) | 22.5% (51/227) | 27.9% (2229/7990) | 33.8% (3579/10577) | 34.4% (1802/5239) | <0.001 |
| Prior peripheral vascular disease | 3.4% (864/25384) | 11.9% (29/244) | 4.1% (347/8488) | 3.0% (335/11132) | 2.8% (153/5520) | <0.001 |
| Number of days hospitalized | 4.0 [3.0, 5.0] | 4.0 [3.0, 8.0] | 4.0 [3.0, 6.0] | 4.0 [3.0, 5.0] | 4.0 [3.0, 5.0] | <0.001 |
| Acute occlusion | 60.9% (15460/25384) | 54.1% (132/244) | 60.4% (5129/8488) | 61.4% (6830/11132) | 61.0% (3369/5520) | 0.46 |
| Intraprocedural thrombosis | 19.7% (4988/25376) | 16.4% (40/244) | 19.0% (1609/8487) | 20.5% (2278/11128) | 19.2% (1061/5517) | 0.05 |
| Successful procedure | 95.3% (24017/25194) | 91.5% (215/235) | 95.2% (8016/8420) | 95.8% (10595/11061) | 94.8% (5191/5478) | 0.85 |
| Aspirin pre-cath lab | 95.3% (24017/25194) | 91.5% (215/235) | 95.2% (8016/8420) | 95.8% (10595/11061) | 94.8% (5191/5478) | 0.78 |
| Clopidogrel pre-cath lab | 19.0% (4814/25366) | 18.9% (46/244) | 19.3% (1638/8484) | 19.4% (2153/11126) | 17.7% (977/5512) | 0.12 |
| Ticagrelor pre-cath lab | 58.1% (13800/23736) | 54.6% (125/229) | 56.9% (4515/7941) | 58.5% (6077/10385) | 59.5% (3083/5181) | 0.19 |
| Prasugrel pre-cath lab | 4.1% (1032/25368) | 2.9% (7/244) | 4.0% (342/8484) | 4.2% (465/11127) | 4.0% (218/5513) | 0.70 |
| Warfarin pre-cath lab | 2.1% (526/25368) | 2.0% (5/244) | 2.2% (185/8484) | 2.0% (223/11127) | 2.0% (113/5513) | 0.86 |

\*Data are presented as mean ± SD, percentages (numerator/denominator) or medians [interquartile ranges]

For the first primary endpoint, participants were followed until death or for 30 days; and for the secondary primary end point, participants, were followed until death or up to 1 year. At these time points the SCAAR and Cause of Death Register were linked. Death is presented as a cumulative incidence function and comparison between groups was performed with the log rank test. Persons who emigrated or were alive at the end of the 1- or 12-month follow-up, respectively, were treated as censored observations. Death within the first 30 days was included as an event in the analysis for outcome at one-year follow-up, since we felt that that the risk for bias was low and, therefore, a landmark analysis redundant. Furthermore, since there were no competing events, it was not necessary to take this into account in the statistical analysis.

To evaluate the association between BMI categories and mortality, univariate and multivariable-adjusted hazard ratios (HR) were calculated using Cox proportional-hazards regression models. We performed a primary investigation employing a complete case analysis. Thereafter, to handle missing data, we performed a secondary examination after multiple imputation missing pattern [MIMP] with the missing at random (MAR) assumption. The first model was unadjusted; the second model was adjusted for age; the third model was adjusted for age and sex; and the fourth and final model was adjusted for all co-variates listed in **Table 1**. The reference group was defined as the normal nutritional category as defined by the WHO corresponding to a BMI of 18.5–24.99 kg/m^2^.

All models were specified prior to conducting analyses and adjusted for preselected baseline risk factors considered of importance for the outcome. Further, the causal directed acyclic graph approach (DAGs) was applied when adjusting for confounding. An adjusted mortality analysis for the first 30-days was also considered but, due to few events, we concluded that the study was underpowered for this. Penalized spline regression was applied to study relationship between BMI as a continuous variable and all-cause mortality. The likelihood ratio test was used to examine the consistency of the association between BMI categories and mortality in the following subgroups defined by baseline characteristics: males vs females, age > 65 years vs age ≤ 65 years; presence or absence of diabetes; and smokers vs non-smokers. All statistical tests were two-tailed and P-values of <0.05 were considered statistically significant.

## Results

### Patient characteristics

Between January 1^st^ 2011 and May 31^st^ 2018, a total of 25,384 patients underwent coronary artery catheterization for ST-elevation myocardial infarction at 29 PCI centers in Sweden. Among these, a total of 1304 (5.1%) died within 30 days of PCI and 5,529 (21.8%) died within one year after the intervention. Baseline characteristics for the total study group, and for different BMI categories separately, are presented in **Table 1**. People with obesity tended to be younger and have a more adverse cardiovascular risk factor profile with higher frequencies of hypertension, hyperlipidemia and diabetes as compared with those in other BMI categories, but were less often smokers than those who were underweight. Underweight patients were more often females, smoked more frequently and had a higher prevalence of chronic obstructive pulmonary disease (COPD). The angiographic burden of coronary artery disease and number of days hospitalized were similar among patients with different BMI categories.

### Unadjusted 30-day and 1-year mortality in different BMI classes

Unadjusted *30-day* all-cause mortality for different BMI categories are presented with cumulative incidence curves in **Figure 1**. Patients who were underweight had the highest 30-day mortality (13.3%), followed by patients with normal weight (6.6%). Overweight and obese patients had somewhat lower cumulative mortality (4.3% and 4.2%, respectively). The overall log-rank p-value was <0.001.

**Figure 1.** Thirty-day all-cause mortality after PCI in STEMI for different BMI categories

Unadjusted *1-year* all-cause mortality for different BMI categories is presented with cumulative incidence curves in **Figure 2**. Again, there was a substantial difference in mortality between different BMI classes with the highest risk in the underweight group (23.3%) followed by those with normal weight (11.3%). Patients who were overweight or obese had a lower 1-year mortality risk (7.3% and 6.9%. respectively). The overall log-rank p-value was < 0.001.

**Figure 2.** One-year all-cause mortality after PCI in STEMI for different BMI categories

### Adjusted 1-year mortality in different BMI categories

**Figure 3** presents a Forest plot of hazard ratios displayed in log-10 scale for *1-year* all-cause mortality in different BMI categories using the normal weight population as a reference group. People with underweight had the highest mortality compared with people of normal weight in both unadjusted and multi-adjusted analysis. In contrast, patients with overweight had a lower mortality-risk compared with normal weight in both unadjusted and multi-adjusted analysis. Patients with obesity had a lower risk for death than normal weight subjects in unadjusted analysis, but this difference became non-significant in the multi-adjusted analysis considering age, sex and other covariates. Still the HR did not differ much from the overweight group and it is possible that he lack of significance is a type II statistical error.

**Figure 3.** Unadjusted and adjusted risk for mortality (95% CI) in patients with STEMI using log-10 scale for the X axis

In **Figure 4** we examined the association between BMI as a continuous variable and unadjusted and adjusted all-cause mortality using fractional polynomial Cox regression. In models adjusted for age and sex, the curves were U-shaped with a BMI/risk nadir between 25.0-29.9 kg/m^2^. In the unadjusted and fully adjusted model the right side of the curve flattened with wide confidence intervals. In an interaction analysis, the relationship between BMI categories and risk of death was similar in subgroups of selected baseline characteristics (**Table 2**).

**Table 2.** Hazard ratios for the risk of mortality in subgroups.

| Subgroups | BMI category | HR (95% CI) | P-value for interaction |
| --- | --- | --- | --- |
| Sex |  |  |  |
| Male | Underweight | 2.074 (1.272-3.383) | 0.68 |
|  | Overweight | 0.873 (0.756-1.008) |  |
|  | Obese | 0.890 (0.730-1.084) |  |
| Female | Underweight | 1.394 (0.917-2.119) |  |
|  | Overweight | 0.866 (0.721-1.039) |  |
|  | Obese | 0.864 (0.691-1.082) |  |
| Age |  |  |  |
| Age > 65 | Underweight | 1.595 (1.140-2.230) | 0.38 |
|  | Overweight | 0.896 (0.794-1.011) |  |
|  | Obese | 0.911 (0.771-1.076) |  |
| Age ≤ 65 | Underweight | 1.964 (0.714-5.406) |  |
|  | Overweight | 0.687 (0.495-0.952) |  |
|  | Obese | 0.709 (0.492-1.023) |  |
| Diabetes mellitus |  |  |  |
| Diabetic | Underweight | 2.591 (1.051-6.385) | 0.62 |
|  | Overweight | 0.937 (0.736-1.194) |  |
|  | Obese | 0.877 (0.674-1.143) |  |
| Non-diabetic | Underweight | 1.535 (1.092-2.157) |  |
|  | Overweight | 0.851 (0.748-0.968) |  |
|  | Obese | 0.898 (0.745-1.082) |  |
| Smoking |  |  |  |
| Current smoker | Underweight | 1.766 (1.017-3.066) | 0.67 |
|  | Overweight | 0.782 (0.611-1.001) |  |
|  | Obese | 0.917 (0.674-1.249) |  |
| Non-smoker | Underweight | 1.550 (1.049-2.291) |  |
|  | Overweight | 0.893 (0.786-1.015) |  |
|  | Obese | 0.871 (0.733-1.034) |  |

**Figure 4.** Unadjusted and adjusted fractional polynomial Cox proportional hazards regression (95% CI, shaded area) with continuous risk relationship between BMI and all-cause mortality after PCI treatment for STEMI

## Discussion

In the present study, unadjusted statistical analysis showed that people with obesity (BMI ≥30.0 kg/m^2^) had the lowest *30-day* and *1-year* all-cause mortality after coronary intervention due to STEMI, when compared with the subjects of normal weight (BMI 18.5-24.99 kg/m2). Thus, in these analyses our results were in line with the obesity paradox. However, after adjustment for age and sex, the effect of obesity on mortality did not differ from that observed in people of normal weight. Instead, when age and sex had been considered, people with overweight (BMI 25-29.9 kg/m2) had the lowest mortality. In the one-year model adjusted for all covariates, people with overweight still had the lowest death rate, whereas the risk in people with obesity did not differ from that of those with normal weight. However, increasing BMI as a continuous variable in the fully adjusted one-year analysis, the risk curve flattened out with a wide confidence interval, making it difficult to interpret. Of notice, people with obesity do have a lower mortality in unadjusted analysis and did not display a higher mortality than those of normal weight in the fully adjusted model. On the other hand, people who were underweight (BMI <18.5 kg/m2) had a mortality that was substantially higher than that observed for other BMI classes.

Previous publications, including a report from the Framingham heart study (19), results from the Canadian APPROACH register (20) and data from the SCAAR register (12) showed that in patients with established coronary artery disease, the lowest adjusted risk for mortality reached a nadir around a BMI of 35 kg/m^2^. Furthermore, a meta-analysis by Wang and et al. (21) found lower all-cause mortality after myocardial infarction in a pooled group of overweight and obese subjects compared with people of normal weight. In contrast Shahim and coworkers (22) found no relationship between BMI and myocardial infarction size, or 1-year rates of death or heart failure hospitalization, in a meta-analysis based on 2,238 patients undergoing PCI.

The present study differed from the publications cited above regarding several aspects. Our patient population was considerably more homogenous compared with that in previous studies. The heterogenicity of previous study populations involved inclusion of patients undergoing PCI for STEMI, also subjects with NSTEMI treated with PCI or medication alone and in some cases management with coronary artery bypass grafting. Furthermore, in some of these studies patients with underweight were excluded and adjustment for covariables were not performed as they were not available. It is not unlikely that these differences may explain the main disparity between previous reports and the present study, which showed that the lowest risk for one-year all-cause mortality after adjusting for covariates was observed among overweight patients (BMI-range 25.0-29.9 kg/m^2^). Our findings are more in line with those of Flegal et al (23) who performed a metanalysis including 2.9 million people and observed that the overweight group had a trend for better survival as compared with those who were obese.

In the present study underweight individuals (BMI <18.5 kg/m^2^) displayed the highest risk for all-cause 30-day and 1-year mortality post-PCI in both unadjusted and adjusted statistical analyses. The separation between the mortality curves for the underweight group, as compared with other BMI categories, occurred early after PCI and, showed, thereafter, a steeper upward slope during one-year follow-up. We speculate that underweight patients may have an underlying pathophysiology that may generate a larger STEMI, more complications or impaired recoverability than subjects with a higher BMI. Also, previous epidemiological studies have observed a U-shaped relationship between BMI and all-cause mortality in the general population (24, 25). This has mainly been attributed to smoking (26), respiratory illnesses (27) and other underlying diseases (28). In the present study underweight patients were more often smokers and had a higher prevalence of chronic obstructive pulmonary disease (COPD). Thus, excessive mortality in underweight patients following PCI for STEMI could also be related to a higher occurrence of underlying co-morbidities.

The relationship between categories of BMI and outcome was consistent across all subgroups studied for selected baseline characteristics. Hence, there was no difference between the two sexes, those with age below or above 65 years, those with or without diabetes mellitus, or those who smoked and those who did not.

In a review from 2017, Lavie et al. (29) have listed several possible reasons or biases for the obesity paradox including younger patients, fewer smokers, better energy reserves, increased muscle mass and reverse epidemiology due to frailty and cachexia in patients that are leaner, apart from other and unknown confounders. Of notice is that BMI as a measure of obesity does not differ between fat, muscle mass and skeletal weight (30). It is highly probable that an augmented muscle mass may act as a protective factor with respect to outcome after coronary interventions (31). Lavie et al studied patients with stable coronary artery disease and found that those with a higher lean body mass had better survival irrespective of their degree of fatness (32). Thus, the importance of muscle mass as an explanatory mechanism of the obesity paradox has probably been underestimated. Also, cardiorespiratory fitness is an important variable that may greatly influence the relationship between obesity and survival after PCI (33).

### Strengths and limitations

The strength of our study includes a large sample size of real-world data and a homogenous group of patients treated with PCI for ST-elevation myocardial infarction. The main limitation is the observational nature of the study, which precludes us from making any causal inferences. As only surviving hospitalized patients are included, the possibility of selection bias, residual confounding and survival bias cannot be ruled out. BMI as a measure of obesity, has its limitation as a measure of obesity since it does not distinguish between fat, muscle mass and skeletal weight. Neither waist circumference nor other measures of abdominal fatness were not available in the SCAAR register and, therefore, we were not able into account the distribution of body fat in our analyses. The role of unintentional weight loss was not controlled and cause-specific mortality data was not studied. Data on death was collected by crosslinking the SCAAR with the Swedish Cause of Death Register, which is a high quality virtually complete register of all deaths in Sweden since 1952, but is not adjudicated to establish cardiac versus non-cardiac causes of death.

### Conclusions

Although people with obesity displayed lower mortality after treatment with PCI for STEMI as compared with a reference group with normal weight, the two groups showed similar outcomes after relevant covariates were considered. Assessed against the referent group, overweight patients showed the lowest 30-day and one-year adjusted mortality risk, and underweight individuals the highest. We speculate that the amount of muscle mass and cardiorespiratory fitness may affect the relationship between BMI and outcome in patients with coronary artery disease.

## Data Availability

On request the data is available for examination

## Contributorship Statement

Shabbar Jamly, MD, PhD. Conceptualization, investigation, methodology, project administration, visualization, supervision, validation and writing original draft. Review and editing. Overall content guarantor.

Björn Redfors MD, PhD. Conceptualization, data curation, formal analysis and software.

Lena Carlsson, MD, PhD. Writing -review and editing

Elmir Omerovic, MD, PhD. Conceptualization, data curation, writing-review and editing.

Kristjan Karason, M.D. Ph.D. Funding acquisition, writing -review and editing. Overall content guarantor.

